# Trade Study of Population Age Groups for a Clinical Trial in Attention Deficit Hyperactivity Disorder

**DOI:** 10.1101/19009639

**Authors:** Dalton B. Lovinsand, Robert A. Lodder

## Abstract

ADHD is a neurodevelopmental disorder characterized by difficulty paying attention, impulsivity, and hyperactivity that is treated by an array of different modalities in order to decrease functional impairment. Despite the variety of available pharmacologic treatment options and their widespread use, there still remains a strong need to create innovative therapies. Game Based Therapy (GBT) is a type of cognitive behavioral therapy that seeks to increase the efficacy of currently available pharmacological treatments through improving cognitive deficits and reducing the current dose of such treatments. At the University of Kentucky, a clinical trial was implemented to assess the effects of combining stimulant pharmacotherapy with the popular video game Minecraft as Game Based Therapy on ADHD symptoms in children ages 10-15. However, the low subject recruitment played a large role in the trial failing to produce conclusive data from that site.

This trade study serves to address the difficulties of subject recruitment and determine an optimal population and location for future adoption of the clinical trial. Through statistical analysis of state, national, and survey data it was found that in Lexington, Kentucky, 18-21 year olds are the optimal population for future GBT with pharmacotherapy clinical trials.

## INTRODUCTION

Attention deficit hyperactivity disorder (ADHD) is a neurodevelopmental disorder characterized by difficulty paying attention, impulsivity, and hyperactivity^1^ and is the most commonly diagnosed behavioral disorder in children.^2^ ADHD was originally defined in children but is now recognized to persist into adulthood for some patients. Despite this recognition, adult ADHD remains underdiagnosed.^3^ A study using a nationally representative sample of adults in the United States estimated that the prevalence of ADHD in adults was 4.4%.^4^

Importantly, symptom presentation changes over a lifetime in individuals with ADHD and can be associated with different profiles of functional impairment. Symptoms of hyperactivity, impulsivity, and inattention during childhood often result in disruptive behavior at home and in academic impairment at school. In adulthood, symptoms further evolve such that hyperactivity decreases or morphs into more purposeful activity or inner restlessness, whereas inattention, disorganization, and impulsivity remain, which can lead to functional difficulties in home, social, and work settings.^5,6^

Appropriate management of adult patients with ADHD is multimodal. Psychoeducation, counseling, supportive problem-directed therapy, behavioral intervention, coaching, cognitive remediation, and couples and family therapy are useful adjuncts to medication management. Concurrent supportive psychosocial treatment or polypharmacy may be useful in treating the adult with comorbid ADHD.^3^

An array of nonpharmacologic and pharmacologic treatments are available for the treatment of ADHD in adults. Nonpharmacologic therapies, such as cognitive behavioral therapy (CBT), dialectical behavioral therapy (DBT), physical exercise, and mindfulness awareness practice, have all been shown to confer benefits in adults with ADHD.^3^ CBT is a model that combines higher-level organization and planning, behavioral skills training, and cognitive restructuring. CBT has been used extensively as a stand-alone therapy and in combination with pharmacotherapy in the treatment of ADHD in adults.^7^ Pharmacologic options include psychostimulants, which have demonstrated the greatest levels of efficacy for treating the core symptoms of ADHD in adults, and nonstimulants, which can be considered when there are concerns related to response, tolerability, or safety with psychostimulants. Despite the variety of available pharmacologic treatment options and their widespread use, there still remains a strong need to create innovative therapies.^8^

Unfortunately, the benefits that do result from stimulant use do not persist after discontinuation and patients with ADHD still suffer from adverse long-term outcomes such as poor academic performance, drug addiction, and criminal behavior to a much greater degree than non-ADHD subjects despite optimal therapy.^9^

The advantages of combining pharmaceutical therapy with behavioral therapy include lower cost, as personnel time with patients is reduced, and reduced doses of pharmaceuticals (50-75%). Additionally, the benefits of behavior therapy are more likely to carry forward once treatment has stopped.^10^

The possibility of a synergistic effect from combining pharmacological therapy with Game Based Therapy (GBT) for use in ADHD treatment stems from being able to increase the efficacy of currently available pharmacological treatments through improving cognitive deficits and having the ability to reduce the current dose of such treatments. The suggested mechanism for GBT improving ADHD symptoms is that improves the cognitive processes that underlie them. Executive function, a potential intervention target, is a set of top-down mental processes that regulate distracting influences and automatic, unproductive behaviors.^11^ As a result, targeting executive function may allow patients to have better self-control.

Another advantage of utilizing video games for combination therapy is the fact that there is a high volume of scenarios upon which to strengthen patients skills. Different scenarios each day throughout treatment periods reduce the training effect and allow the patient to broaden complex thinking skills instead of focusing on memorizing how to complete a specific task.

One study produced by Akili Interactive Labs, Inc. compared the cognitive functions’ of neuro-typical children/those that lacked a diagnosis of ADHD with children that were diagnosed with ADHD. Akili uses a technology entitled EVO to evaluate cognitive interference and the potential maintenance of clinical benefit produced from game based therapy in children ages 8-12 with ADHD. EVO is used to assess reaction time, multitasking ability, and sustained effects of game-based therapy after the end of treatment periods. The study found that there was a significant improvement in the reaction time for the children with ADHD, and lacked a statistically significant result for the neurotypical children.

Minecraft is an online video game containing a virtual land where users can create their own worlds and experiences, using building blocks, resources discovered on the site and their own creativity. Minecraft requires its users to apply problem solving, planning, and organizational skills for creative building and exploration.^12^ It offers many levels of difficulty, which allows for adaptation to users with a wide range of cognitive abilities as well as potential for incremental progressions in difficulty as the user’s performance improves.

Our previous clinical study utilized the popular online video game, Minecraft, to evaluate its potential for improving executive function in children, ages 10 to 15, with ADHD. The personalized medicine approach included an individualized treatment regimen consisting of an initial stimulant dose recommendation and schedule of Minecraft activities which were determined by the initial ADHD assessment results of new patients.^13^ The study was designed to create novel training activities that would address both the underlying core executive functions (working memory, inhibition, and cognitive flexibility), as well as executive function skills utilized in school and work settings (planning, time management, self-regulation, sustained attention, organization, goal-directed persistence, and metacognition). One of the overall goals of the trial was to develop a better understanding of the relationship between NICHQ Vanderbilt Assessment scores (an assessment that measures ADHD symptoms as well as the patient’s function in everyday life), game aspects of Minecraft, and executive function.

This paper will serve as a trade study^14^ to collect and compare relevant information regarding two different age groups. A trade study (also known as a trade-off study) serves to analyze the differentiating factors between two or more systems and identify the most balanced technical solution. Trade studies utilize both quantitative and qualitative measures when determining which solution will produce the optimal outcome. They examine critical factors such as system cost, reliability, maintainability, logistic support, growth potential, and more to improve systematic decisions. For example, if the challenge for an automotive company is to design a vehicle that will bring in the most profit for the company, a trade study can be performed. This allows automotive teams to look into different manufacturing materials, locations for production, fuel-efficiency systems, etc. so that they can create different vehicles, then identify the one with the best profit margin. For a clinical trial, there are many outcome-influencing factors as well. For example, a disease may be location specific, like Ebola, so if an organization were to do a clinical trial in a low-prevalence area, the results of the study may not reflect the entire population.

The initial study^13^ sought to observe the effects of playing Minecraft on executive function in pediatric patients, ages 10-15, with ADHD. The exploratory study used specially designed technological activities to train executive function, working memory, and restraint within the environment of the popular computer game Minecraft. The study attempted to determine whether there is a functional relationship between NICHQ scores, game aspects of Minecraft, and executive function. The primary measures of this study were the changes in NICHQ scores, as measured by a NICHQ questionnaire before and after 4 weeks of therapeutic gaming while simultaneously taking a stimulant type ADHD medication. All study participants were recruited within UK Healthcare in Lexington, Kentucky from October of 2016 to June of 2018.

The trial studying the 10-15 year old adolescent population in Lexington, Kentucky was launched to determine the possibility of synergistic effects from GBT and pharmaceutical therapy for ADHD.^13^ However, the trial failed to recruit a population large enough to be able to make an accurate scientific conclusion. One major obstacle the study encountered was that you cannot recruit in public schools unless you invest in getting permission. However, university students are readily available, thus it may be easier to conduct a study in the 18-21 year old college student population.

This trade study will address specific risks, benefits, challenges, and future clinical research directions associated with combining pharmacological and GBT for these two different populations. The paper will also reveal the value of reproducing the initial study with the 18-21 year old population as well as address the reasons for low recruitment in the past. In order for the future study to be executed we must meet requirements that include a minimum 16 subject population that is recruitable within two months and an IRB approved population.

For the two different study groups we will assess 13 different categories that we believe have a significant effect on subject recruitability and/or IRB approval. These categories and the values associated with them will serve as evidence for determining statistical differences between the two age groups. The 13 categories are as follows: transportation, the IRB parent/guardian approval requirement for those under the age of 18, financial need, ADHD rate, population density, population proximity, video game experience, life experience, student/teacher relationship strength as determined by class size, medication therapeutic range, obesity rate, sleeping disorder rate, and anxiety disorder rate. The descriptions of each category are included in our scoresheet_here.

### STATISTICAL ANALYSIS VALUES ARE IN **[BRACKETS]**

| Category | Adolescents (10-15y) | College Students (18-21y) |
| --- | --- | --- |
| **Transportation<sup>#</sup>** | **[10]<sup>1,2</sup>**<br>10% availability<br>Biking/walking distance,<br>local buses, taxi services,<br>dependent upon parents<br>transportation or parent<br>approval of transportation<br>form. | **[100]<sup>2,3</sup>**<br>Independent on others for<br>transportation.<br>Walking/biking distance,<br>bus routes,<br>self-owned vehicle. |
| **Parent/Guardian**<br>**Approval**<br>**Requirement<sup>#</sup>** | **[0]<sup>4</sup>**<br>Required | **[100]<sup>4</sup>**<br>Not required |
| **Financial Need Rate** | **[17.8]<sup>5</sup>**<br>17.8% are below the poverty<br>level | **[70.0]<sup>6</sup>**<br>70% of students have debt<br>after graduation.<br>On average, \$29,800 owed<br>after graduation. |
| **Population Density**<br>**(Lexington, KY)** | **[21.9]<sup>7</sup>**<br>n=16,175<br>Male: 8,283<br>Female: 7,892<br>Per square km<br>(16,175/739km<sup>2</sup>)<br>= 21.9 | **[23.9]<sup>7</sup>**<br>n=17,671 (18-21)<br>Male: 8,780<br>Female: 8,891<br>Per square km = 17671/739 =<br>[23.9] |
| **Proximity to UK Healthcare\*** | **[4.0]<sup>8</sup>**<br>3.97 miles<br>Average distances from 5 middle and 5 high schools in Lexington, KY to UKHC | **[0.7]<sup>9</sup>**<br>0.7 miles between the UK Main Building and UKHC |
| **ADHD Rates** | **[6.1]<sup>10</sup>**<br>Of studied 4-17 year olds, 6.1% take medication for ADHD in US | **[5.0]<sup>11</sup>**<br>2-8% of college students across the US |
| **Video Game Experience<sup>#</sup>** | **[.5]**<br>On average this sample will have less experience due to age and video game comprehension difference. | **[1.0]**<br>On average this sample will have approximately double the amount of time based on the amount of years of video game playing capability. |
| **Life Experience** | **[12.5]**<br>Age (10-15) mean | **[19.5]**<br>Age (18-21) mean |
| **Student/Teacher Relationship Strength as Determined by Class Size\*** | **[19.2]<sup>12</sup>**<br>average of 5 major high schools in Lexington | **[17.0]<sup>13,14</sup>**<br>17:1 Student:faculty ratio at the University of Kentucky |
| **Obesity Risk\*** | **[15.8]<sup>15</sup>**<br>15.8% of students aged 10-17 had obesity | **[10.2]<sup>16</sup>**<br>“While obesity rates for college-age students remains relatively low (males 10.8%, females 9.6%)” |
| **Therapeutic dosing range\*\*** | **[25]<sup>17</sup>**<br>Most common stimulant for UKHC - Dextroamphetamine<br>For patients ages 6 and over, the starting dosage is typically 5 mg once or twice a day.<br>Mayo Clinic states the daily dose is typically not over 30. (5mg-30mg) (30-5=25) | **[35]<sup>18</sup>**<br>Dextroamphetamine - Usual dose ranges from 5 to 40 mg per day in 1 to 3 divided doses. Subsequent doses may be given at 4 to 6 hours intervals<br>(5-40)<br>(40-5=35) |
| **Sleeping disorder rate\*** | [3.7]<sup>19</sup><br>Across all ages, 3.7% of youths had an ICD-9 diagnosis for a sleep disorder. 100,000+ sample size | [27]<sup>20</sup><br>Twenty-seven percent of students were at risk for at least one sleep disorder. 1800+ sample size in southeastern college. |
| **Anxiety Disorder Rate\*** | [25.1]<sup>21</sup><br>Anxiety disorders affect 25.1% of children between 13 and 18 years old | [21.9]<sup>22</sup><br>21.9 percent of students said that within the last 12 months, anxiety had affected their academic performance. |
| **Population Density**<br>(For NYC, **CHC**, ATL) | NYC - 749,424/784km<sup>2</sup> = [955.9]<sup>23</sup><br>**CHC - 100,579/591km<sup>2</sup> = [170.2]<sup>24</sup>**<br>ATL - 11,886/343km<sup>2</sup> = [34.7]<sup>25</sup> | NYC - 485,646/784km<sup>2</sup> = [619.4]<sup>23</sup><br>**CHC - 69,268/591km<sup>2</sup> = [116.9]<sup>24</sup>**<br>ATL - 9,241/343km<sup>2</sup> = [26.9]<sup>25</sup> |
| **Location Proximity\***<br>(For NYC, **CHC**, ATL) | Distance from local NYC High Schools to NYU Global Center: [.833]<sup>26</sup><br>**Local HS/Middle schools to UChicago Medical Center: [1.13]<sup>28</sup>**<br>Local ATL High schools to Emory University Hospital: [3.97]<sup>30</sup> | Distance from NYU to NYU Medical Center: [0.2]<sup>27</sup><br>**From UChicago to UChicago Medical Center: [0.2]<sup>29</sup>**<br>From Emory University to Emory University Hospital: [0.4]<sup>31</sup> |
| **Student/Teacher Relationship as Determined by Class Size\***<br>(For NYC, **CHC**, ATL) | NYC - [26.4] citywide<sup>32</sup><br>**CHC - [29.1] local HS/MS<sup>34</sup>**<br>ATL - [13] local HS/MS<sup>36</sup> | NYU - [30]<sup>33</sup><br>**UChicago - [15]<sup>35</sup>**<br>Emory University - [25]<sup>37</sup> |

\* = values between age groups were switched for statistical analysis

### A summary of the resulting comparison

To reiterate, in order to compare the 10-15 year old age group to the 18-21 year old age group, thirteen different characteristics that may have an influence on an ADHD GBT clinical trial were identified. These characteristics were then given values for each age group based upon an in-depth literature search that served to reflect approximate demographics for Lexington, Kentucky when obtainable. If Lexington demographics were unavailable, national demographics were used. If national demographics or outside references were unavailable, a heuristic value was given. Heuristic values are like “rules of thumb”, values which are not guaranteed to be the true, real-world value, but are sufficient for immediate goals.^15^ Values that are heuristic are indicated as such within the table above.

Throughout all tables within this study, higher values are designated more valuable to recruitability and IRB approval versus lower values. However, certain category values were switched between age groups due to the higher number being less desirable (e.g. being 10 miles away from the clinical site is less desirable than 5 miles away-so the ‘10 miles away’ group received a value of 5, and vice-versa. The categorical values that were switched between age groups included: location proximity, student/teacher relationship as determined by class size, obesity risk, therapeutic dosing range, sleeping disorder rate, and anxiety disorder rate.

Four experts were recruited to rank the age group characteristics in order of magnitude of effect on subject recruitment and IRB approval influence, with 1 being the least magnitude and 13 being the most. Two of the subjects lacked prior knowledge of the study and were recruited to reduce subjective bias. The results of the survey are shown below in Table 1.

**Table 1.** Survey Responses

| Ranked from 1-13, with 13 being of highest importance. |  |  |  |  |
| --- | --- | --- | --- | --- |
| Respondent #1 DL | Respondent #2 RL | Respondent #3 PF | Respondent #4 CB | AVG |
| 11 | 12 | 12 | 11 | 11.5 |
| 10 | 11 | 13 | 13 | 11.75 |
| 8 | 5 | 10 | 1 | 6 |
| 13 | 13 | 7 | 10 | 10.75 |
| 12 | 10 | 9 | 9 | 10 |
| 9 | 9 | 8 | 8 | 8.5 |
| 6 | 7 | 6 | 7 | 6.5 |
| 4 | 6 | 5 | 3 | 4.5 |
| 7 | 8 | 11 | 12 | 9.5 |
| 1 | 1 | 2 | 5 | 2.25 |
| 5 | 4 | 4 | 2 | 3.75 |
| 2 | 3 | 3 | 4 | 3 |
| 3 | 2 | 1 | 6 | 3 |

In the first statistical analysis shown below in Table 2, each value for both age groups was converted into a z-score, using the mean and standard deviation of the respective age group, allowing the values to be more comparable to each other. Z-scoring allows us to compare two values that are taken from different distributions, reducing leverage effects. The 26 respective z-scores were averaged between the two age groups, resulting in 13 z-score values. The values were then weighted by multiplying the average subjective score for that specific characteristic. These 13 ‘overall score’ values give us a subjective reference on the magnitude of effect on subject recruitment and IRB approval influence for the corresponding characteristics. The higher overall score correlates to a stronger influence that the characteristic will have on the clinical trial.

**Table 2.** Individual Age Group Values and Z-scores

| Column 10-15 age group Mean | Column 10-15 age group SD | Column 18-21 age group Mean | Column 18-21 age group SD | Overall Column Mean | Overall Column SD |
| --- | --- | --- | --- | --- | --- |
| 13.89 | 10.83 | 31.71 | 35.01 | 22.80 | 26.97 |
| Characteristic | 10-15 age group value | z-score 10-15 age group | 18-21 age group value | z-score 18-21 age group | Overall Score |
| Transportation | 10 | -0.36 | 100 | 1.95 | 9.15 |
| Parent/Guardian Approval Needed | 0 | -1.28 | 100 | 1.95 | 3.93 |
| Financial Need | 17.8 | 0.36 | 70 | 1.09 | 4.36 |
| Population Density | 21.9 | 0.74 | 23.9 | -0.22 | 2.77 |
| Proximity to UKHC | 0.7 | -1.22 | 4 | -0.79 | 10.05 |
| ADHD Rates | 6.1 | -0.72 | 5 | -0.76 | 6.30 |
| Video Game Experience | 0.5 | -1.24 | 1 | -0.88 | 6.87 |
| Life Experience | 12.5 | -0.13 | 19.5 | -0.35 | 1.07 |
| Student/Teacher Relationship | 17 | 0.29 | 19.2 | -0.36 | 0.33 |
| Metabolism/Obesity Risk | 10.2 | -0.34 | 15.8 | -0.45 | 0.89 |
| Therapeutic Dosing Range | 35 | 1.95 | 25 | -0.19 | 3.29 |
| Sleeping Disorder Rate | 27 | 1.21 | 3.7 | -0.80 | 0.62 |
| Anxiety Disorder Rate | 21.9 | 0.74 | 25.1 | -0.19 | 0.83 |

In the second statistical analysis shown below in Table 3, the characteristic values were once again z-scored by age, however this time they were z-scored using the combined means and standard deviations of both age groups. The new 26 z-scores were then multiplied by the average of the subjective scores to produce new subjective magnitudes. Once again, the higher the score, the better the impact on the clinical trial.

**Table 3.**
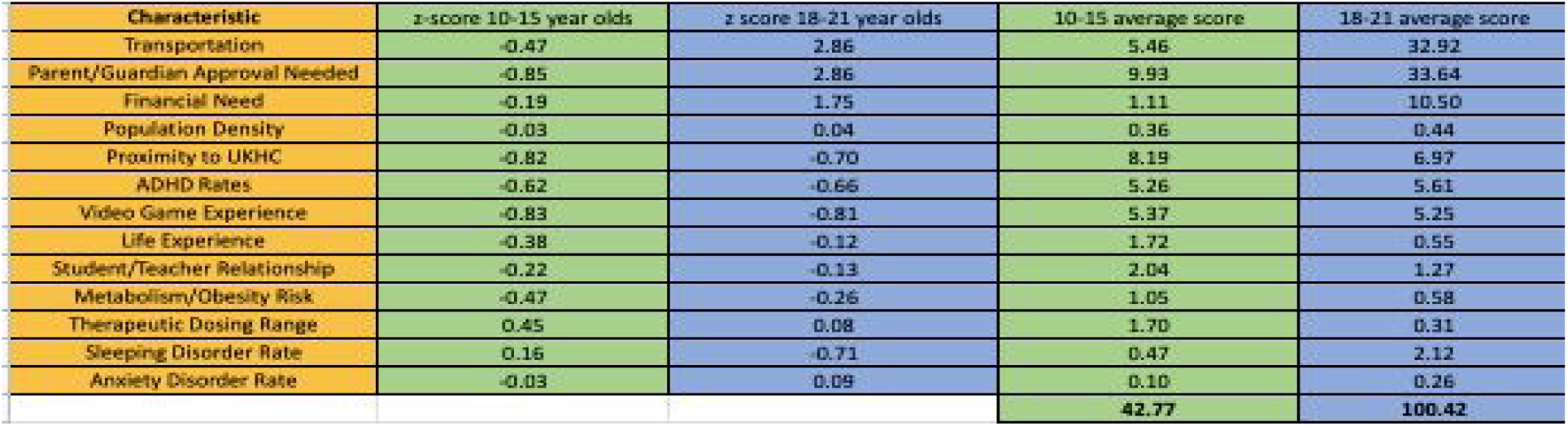
Individual Age Group Values and Z-scores - Combined Mean & Std. Deviation

In the third statistical analysis shown below in Table 4, the higher characteristic values that were deemed optimal for subject recruitability and IRB approval were given a ‘2’ for the age group value, whereas the characteristic values that produced less optimal results for subject recruitability and IRB approval were given a ‘1’ for the respective age group value. Using ‘1’ and ‘2’ for values allows for a simpler version of statistical analysis that focuses less on exact numerical values and more on which age group is preferred for the respective category. The age group values are then multiplied by their respective average subjective score. The resulting individual scores are then totaled by age group. The higher overall score is expected to have the optimal impact on subject recruitability and IRB approval. This categorical analysis shows that regardless of using either exact values from a continuous distribution or categorical preference, the result of the 18-21 year old age group being the optimal population remains the same.

**Table 4.** Simplified Optimal Age Group Values for Subject Recruitment & IRB Approval

| Characteristic | 10-15 age group value | 10-15 score | 18-21 age group value | 18-21 score |
| --- | --- | --- | --- | --- |
| Transportation | 1 | 11.5 | 2 | 23 |
| Parent/Guardian Approval Needed | 1 | 11.75 | 2 | 23.5 |
| Financial Need | 1 | 6 | 2 | 12 |
| Population Density | 1 | 10.75 | 2 | 21.5 |
| Proximity to UKHC | 1 | 10 | 2 | 20 |
| ADHD Rates | 2 | 17 | 1 | 8.5 |
| Video Game Experience | 1 | 6.5 | 2 | 13 |
| Life Experience | 1 | 4.5 | 2 | 9 |
| Student/Teacher Relationship | 2 | 19 | 1 | 9.5 |
| Metabolism/Obesity Risk | 1 | 2.25 | 2 | 4.5 |
| Therapeutic Dosing Range | 2 | 7.5 | 1 | 3.75 |
| Sleeping Disorder Rate | 2 | 6 | 1 | 3 |
| Anxiety Disorder Rate | 1 | 3 | 2 | 6 |
|  |  | **115.75** |  | **157.25** |

In the fourth statistical analysis shown below in Table 5, both of the age group values were divided by the 18-21 year old values to get two respective comparable factors. The new factor values were then multiplied by the corresponding average subjective score to produce concluding values that were totaled for each age group. The higher overall score is expected to have a more optimal expectation for subject recruitability and IRB approval.

**Table 5.** Age Group Factorization Combined with Survey Respondent Scores

| Characteristic | 10-15 age group value | 10-15 age group factor | 10-15 Factor x Respondent Average (Score) | 18-21 age group value | 18-21 age group Factor | 18-21 Factor x Respondent Average (Score) |
| --- | --- | --- | --- | --- | --- | --- |
| Transportation | 10 | 0.1 | 1.15 | 100 | 1 | 11.5 |
| Parent/Guardian Approval Needed | 0 | 0.00 | 0.00 | 100 | 1 | 11.75 |
| Financial Need | 17.8 | 0.25 | 1.53 | 70 | 1 | 6 |
| Population Density | 21.9 | 0.92 | 9.85 | 23.9 | 1 | 10.75 |
| Proximity to UKHC | 0.7 | 0.18 | 1.75 | 4 | 1 | 10 |
| ADHD Rates | 6.1 | 1.22 | 10.37 | 5 | 1 | 8.5 |
| Video Game Experience | 0.5 | 0.5 | 3.25 | 1 | 1 | 6.5 |
| Life Experience | 12.5 | 0.64 | 2.88 | 19.5 | 1 | 4.5 |
| Student/Teacher Relationship | 17 | 0.89 | 8.41 | 19.2 | 1 | 9.5 |
| Metabolism/Obesity Risk | 10.2 | 0.65 | 1.45 | 15.8 | 1 | 2.25 |
| Therapeutic Dosing Range | 35 | 1.40 | 5.25 | 25 | 1 | 3.75 |
| Sleeping Disorder Rate | 27 | 7.30 | 21.89 | 3.7 | 1 | 3 |
| Anxiety Disorder Rate | 21.9 | 0.87 | 2.62 | 25.1 | 1 | 3 |
|  |  |  | **70.40** |  |  | **91** |

The third and fourth statistical analyses were compared below in Table 6.

**Table 6.** Comparison of Data From Tables 4 and 5

|  | Scores for Using 1&2 as factors | Scores using values as factors |
| --- | --- | --- |
| **10-15 group** | **115.75** | **70.40** |
| **18-21 group** | **157.25** | **91** |
| **18-21 / 10-15** | **1.36** | **1.29** |

### #6 - Sensitivity Analysis and Results

In order to test the **subjective** sensitivity of the trade model, the statistical analysis model from Table 3 was used because of its mathematical simplicity. This time however, the average respondent scores were not used, instead results were calculated using individual respondent scores, creating 4 unique respondent results. The respondents included: PharmD/MSPS candidate Dalton Lovins, PharmD/MBA candidate Conner Ball, UK College of Pharmacy pharmaceutical outcomes and policy professor Dr. Patricia Freeman, and UK College of Pharmacy professor and principal investigator for this study, Dr. Robert Lodder. Therefore, the sensitivity test will show how even though different users ranked categories in different positions, the 18-21 year old age group will still succeed in having the highest overall total, making them the optimal group for clinical trial subject recruitability and IRB approval.

### Respondent Results Combined with Simplified Optimal Age Group Values

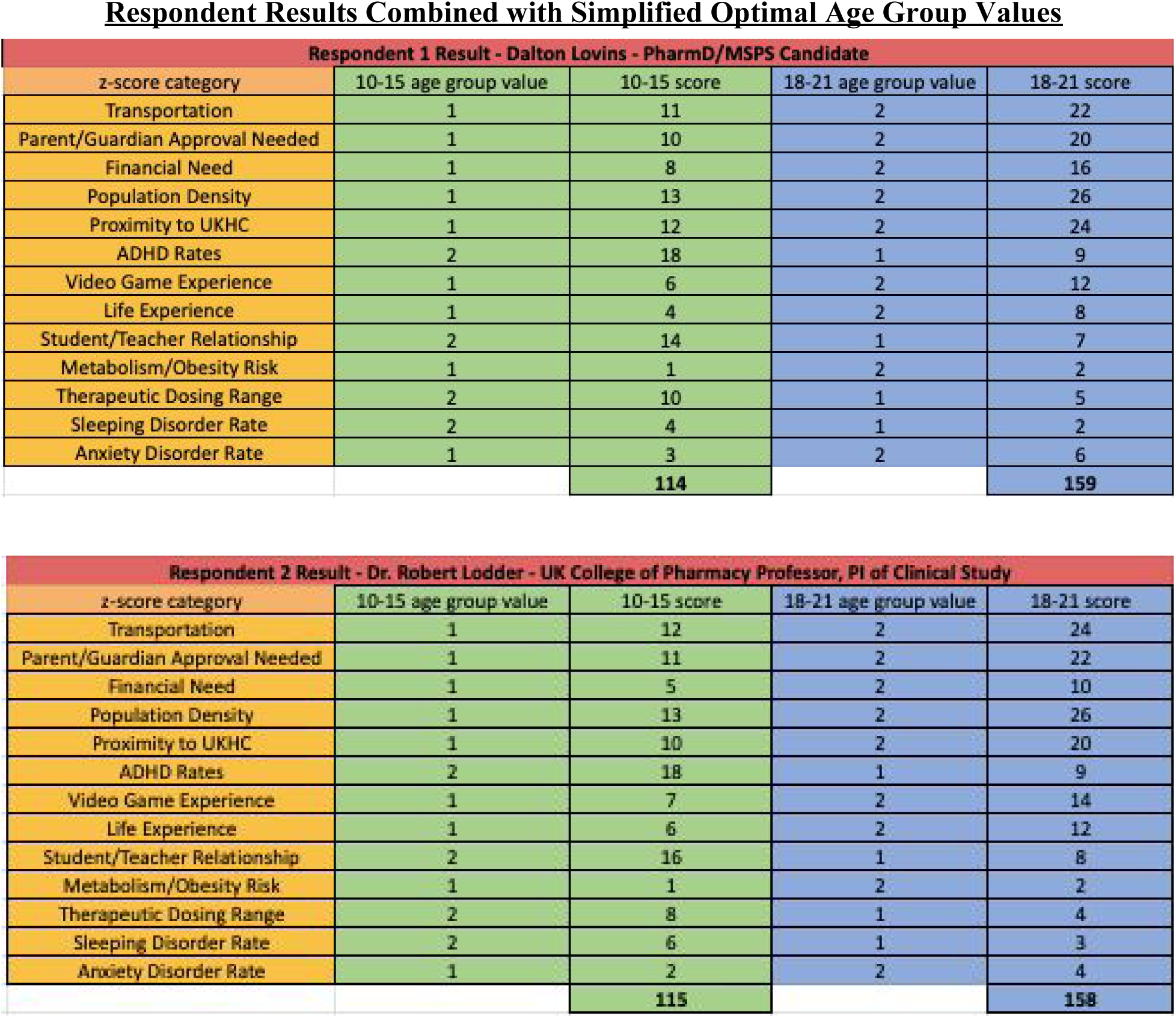
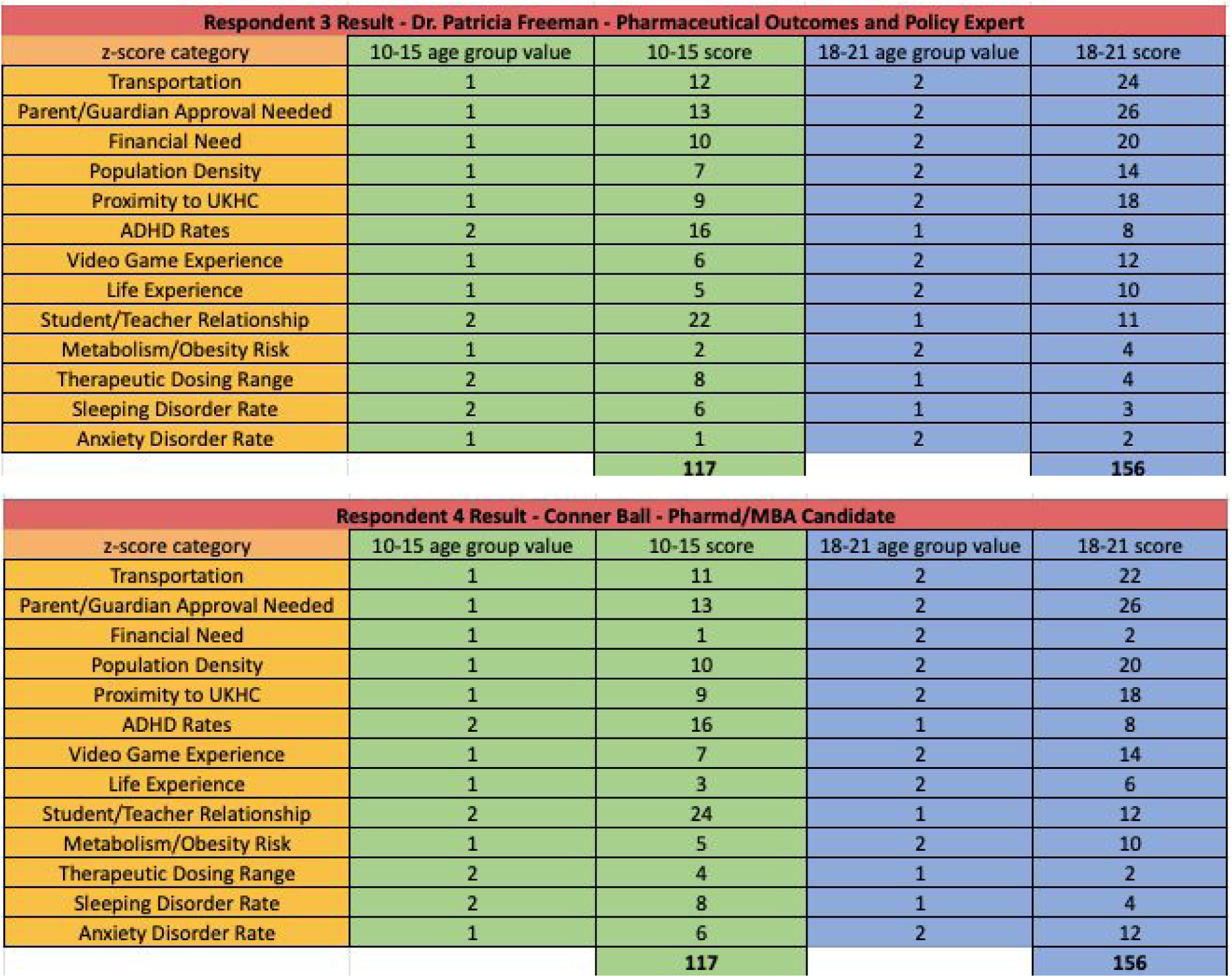

- In order to test the **location** sensitivity of the trade model, we have included results from New York, NY., Chicago, IL., and Atlanta, GA. Even with a population as dense as these 3 major cities, the overall outcome is very similar with the 18-21 year old age group being the optimal treatment group.

### New York City, New York Model

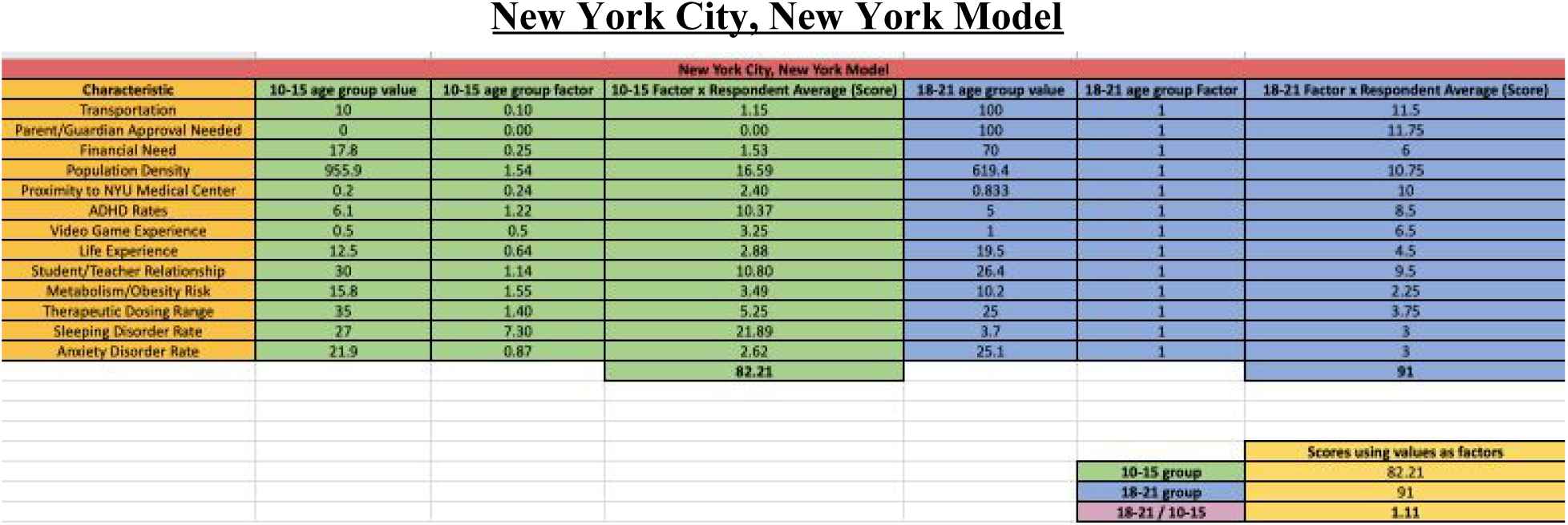

### Chicago, Illinois Model

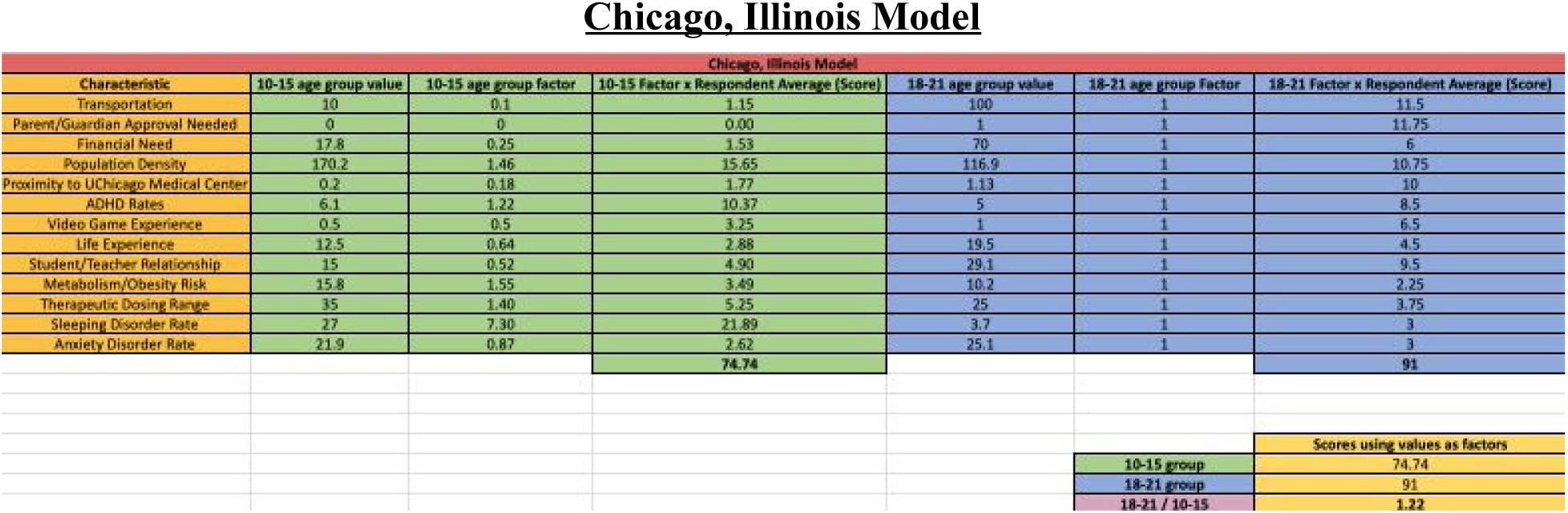

### Atlanta, Georgia Model

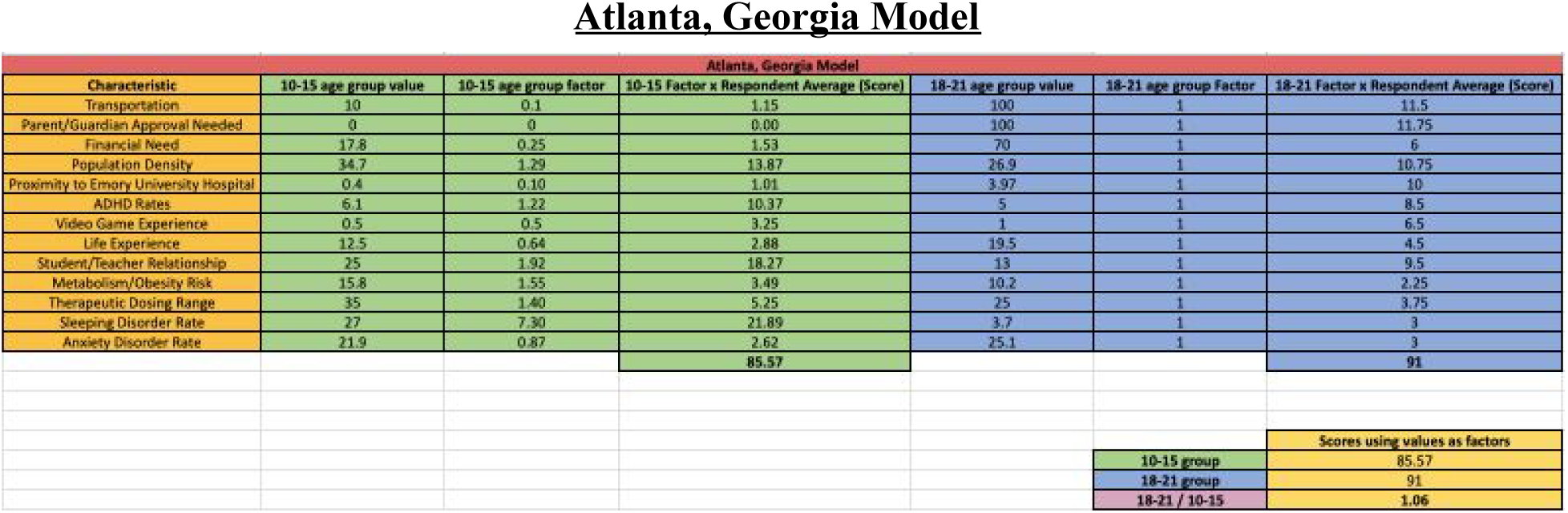

### Conclusion, Adoption, and Further Analysis

The overall conclusion of this study is that the 18-21 year old age group in Lexington, Kentucky is the optimal population for future GBT/ADHD clinical studies.

These data and analyses will be used for the future adoption of the 18-21 year old age group for examining whether GBT has an effect on ADHD symptoms. For the next trial, a maximum of 60 days will be allotted to recruiting at least 16 subjects in order to draw scientific conclusions regarding the relationship between combination Game Based Therapy with pharmacological treatment and ADHD symptoms.

An unexplored option regarding alternative trade studies includes separating the initial survey question, which was, “Rank the age group characteristics in order of magnitude of effect on subject recruitment and IRB approval influence, with 1 being the least magnitude and 13 being the most,” into two separate questions. In turn, one question would ask solely about the magnitude of effect on subject recruitability and the other question would ask solely about the magnitude of effect on IRB approval. This analysis would have been beneficial in comparing the effect on survey responses, and subsequently the outcomes of similarly structured statistical analyses.

## Data Availability

All relevant data files are linked to this manuscript.

https://drive.google.com/file/d/1TYN5v3nJj1ZHGVOadCtib-oFC4p-adZm/view?usp=sharing

## Supplementary Material

A special thanks to those who participated in the clinical trial characteristics ranking. Dr. Lodder is the principal investigator for the initial GBT trial for 10-15 year olds and has been constantly utilized for consults regarding this paper’s purpose, execution, and reviewal. Dr. Patricia Freeman is a current pharmaceutical outcomes and policy professor at the University of Kentucky College of Pharmacy. Conner Ball is a current MBA candidate at the University of Kentucky.

## Acknowledgements

Dr. Robert Lodder - UKCOP Professor

Dr. Patricia Freeman - UKCOP Professor

Conner Ball - PharmD/MBA candidate 2021

Dr. Penni Black - UKCOP Professor

## Statistical Analysis Chart References

Transportation References:

1. Biking: Survey: 100 Million Americans Bike Each Year, But Few Make it a Habit. https://usa.streetsblog.org/2015/03/04/survey-100-million-americans-bike-each-year-but-few-make-it-a-habit/
2. Graduated Driver Licensing Program. Drive.KY.gov. https://drive.ky.gov/driver-licensing/Pages/Graduated-Driver-Licensing-Program.aspx
3. University of KY Campus bus routes https://www.uky.edu/transportation/bus/campusshuttles

Parental Approval Requirement:

4. Field, MJ. Ethical Conduct of Clinical Research involving Children. Institute of Medicine (US) Committee on Clinical Research Involving Children.https://www.ncbi.nlm.nih.gov/books/NBK25560/

Financial Need Rate:

5. Data Tables and Tools ACS. https://www.census.gov/acs/www/data/data-tables-and-tools/
6. A Look at the Shocking Student Loan Debt for 2019. StudentLoanHero. Feb. 9, 2019. https://studentloanhero.com/student-loan-debt-statistics/

Population Density for Lexington

7. Current Lexington, KY Population, Demographics, and stats in 2019, 2018. US Census Data. https://suburbanstats.org/population/kentucky/how-many-people-live-in-lexington

Proximity to UKHC

8. Google Maps - Distance from Morton middle, Windburn middle, Southern Middle, Crawford Middle, Tates Creek Middle, Sayre, Henry Clay, Lex Cath, Trinity Christian Academy Upper School, and Paul Dunbar to UK Chandler Hospital https://www.google.com/maps
9. UK Main Building distance to UK Chandler Hospital https://www.google.com/maps

ADHD Rates - Kentucky

10. CDC. 2011 data. https://www.cdc.gov/ncbddd/adhd/stateprofiles/stateprofile_kentucky.pdf
11. Dupaul GJ, Weyandt LL, O’Dell SM, et al. College students with ADHD: Current status and future directions. J Atten Disord. 2009;13:234−250. doi: 10.1177/1087054709340650

Student Teacher Relationship

12. 17. 2019 Largest Public High Schools in Kentucky. https://www.niche.com/k12/search/largest-public-high-schools/s/kentucky/
13. University of Kentucky FAQs - Average Class size http://students.ca.uky.edu/faqs
14. Why Students Don’t Attend Office Hours https://www.teachingprofessor.com/topics/for-those-who-teach/students-dont-attend-office-hours/

Obesity Risk

15. Childhood Obesity Trends - The State of Obesity. https://www.stateofobesity.org/childhood-obesity-trends/
16. Serious Games for Diabetes, Obesity, and Healthy Lifestyle: Using an Alternate Reality Game to Increase Physical Activity and Decrease Obesity Risk in College Students. https://www.ncbi.nlm.nih.gov/pmc/articles/PMC3440154/

Therapeutic Dosing Range

17. https://www.mayoclinic.org/drugs-supplements/dextroamphetamine-and-amphetamine-oral-route/proper-use/drg-20071758
18. Dextroamphetamine. https://www.ncbi.nlm.nih.gov/books/NBK507808/

Sleeping Disorder

19. Prevalence of Diagnosed Sleep Disorders in Pediatric Primary Care Practices. https://www.ncbi.nlm.nih.gov/pmc/articles/PMC3089951/
20. The Prevalence of Sleep Disorders in College Students: Impact on academic Performance. https://www.csus.edu/faculty/M/fred.molitor/docs/Sleep%20and%20Academic%20Performance.pdf

Anxiety Disorder Rate

21. Facts & Statistics - Anxiety and Depression Association of America. https://adaa.org/about-adaa/press-room/facts-statistics
22. Mental Health Matters: Anxiety and Depression - BU Today. http://www.bu.edu/articles/2016/college-students-anxiety-and-depression

Population Density for New York City, NY

23. Current NYC, NY Population, Demographics, and stats in 2019, 2018. https://suburbanstats.org/population/new-york/how-many-people-live-in-new-york

Population density for Chicago, IL

24. Current Chicago IL Population, Demographics, and stats in 2019, 2018. https://suburbanstats.org/population/illinois/how-many-people-live-in-chicago

Population Density for Atlanta, GA

25. Current Atlanta, GA Population, Demographics, and stats in 2019, 2018. https://suburbanstats.org/population/georgia/how-many-people-live-in-atlanta

Proximity to NYU Medical Center

26. Average distance from Manhattan Village Academy, Norman Thomas HS, LREI Lower and Middle School to NYU
27. Distance between NYU to NYU Langone Medical Center

Proximity to UChicago Medical Center

28. Average distance from University of Chicago Charter School, University of Chicago Laboratory Schools, and King College Prep High School to UChicago Medical Center.
29. Distance between UChicago to UChicago Medical Center

Proximity to Emory University Hospital

30. Average distance from Druid Hills High School, Ben Franklin Academy, Samuel Inman Middle School.
31. Distance between Emory University to Emory University Hospital

Student/Teach Relationship for New York, NY

32. New York City citywide average class size https://infohub.nyced.org/docs/default-source/default-document-library/2018-19_november_class_size_report_-_webdeck_-_11-14-18.pdf?Status=Temp&sfvrsn=c46ddc95_2
33. NYU average class size https://www.collegedata.com/college/New-York-University?tab=profile-overview-tab

Student/Teacher Relationship for Chicago, IL

34. Chicago Public Schools average class size https://cps.edu/About_CPS/At-a-glance/Pages/Stats_and_facts.aspx
35. UChicago average class size https://www.collegedata.com/college/University-of-Chicago?tab=profile-overview-tab

Student/Teacher Relationship for Atlanta, GA

36. Atlanta Public Schools average class size https://www.publicschoolreview.com/georgia/atlanta-public-schools/1300120-school-district
37. Emory University average class size http://www.emory.edu/home/academics/faculty/index.html

